# Genes associated with depression and coronary artery disease are enriched for inflammation and cardiomyopathy-associated pathways

**DOI:** 10.1101/2022.10.25.22280854

**Authors:** Kritika Singh, Hyunjoon Lee, Julia M Sealock, Tyne Miller-Flemming, Peter Straub, Nancy J. Cox, Quinn S. Wells, Jordan W. Smoller, Emily C. Hodges, Lea K. Davis

## Abstract

**Background:** Depression and Coronary Artery Disease (CAD) are highly comorbid conditions. Approximately 40% of individuals who have one diagnosis will also develop the other within their lifetime. Prior research indicates that polygenic risk for depression increases the odds of developing CAD even in the absence of clinical depression. However, the specific genes and pathways involved in comorbid depression-CAD remain unknown.

**Results:** We identified genes that are significantly associated with both depression and CAD, and are enriched for pathways involved in inflammation and for previous association with cardiomyopathy. We observed increased rate of prevalent, but not incident, cardiomyopathy cases in individuals with comorbid depression-CAD compared to those with CAD alone in three electronic large health record (EHR) datasets.

**Conclusions:** The results of our study implicate genetically regulated inflammatory mechanisms in depression-CAD. Our results also raise the hypothesis that depression-associated CAD may be enriched for cardiomyopathy.

**Clinical Perspective:**

A. *What’s New?*
  1. Gene associations shared between depression and CAD are enriched for prior association with cardiomyopathy phenotypes.
  2. Cardiomyopathy is significantly more prevalent in individuals with comorbid depression-CAD than in CAD or depression alone.
B. *What are the Clinical Implications?*
  1. Our work suggests that individuals with comorbid depression-CAD may benefit from screening for cardiomyopathy.

## Introduction

Chronic complex diseases such as cardiovascular disease (CVD) are the primary drivers of premature death among individuals with psychiatric disorders. Approximately 17 to 44 percent of patients with coronary artery disease (CAD), the most common type of CVD, also have a diagnosis of major depression (MDD), a common mental health diagnosis^1^. Several studies suggest that these two conditions are biologically related. A diagnosis of major depression is associated with increased 18-month cardiac mortality among CAD patients; and CAD with comorbid MDD reduces lifespan by 15-20 years^2–5^. However, despite the frequency and high mortality rate of comorbid CAD with depression and/or MDD, hereafter referred to as (major) depressive CAD or (m)dCAD, the biological relationship between these conditions remains poorly understood.

Previously, we demonstrated that common genetic risk for depression identified by genome-wide association studies (GWAS) and quantified in the form of polygenic risk scores (PRS), is associated with a diagnosis of CAD and myocardial infarction in a healthcare-based clinical population^6^. High genetic liability to MDD was associated with increased risk of cardiovascular disease even among patients with no history of psychiatric illness and after accounting for cardiovascular disease risk factors^6^. Shared inflammatory processes provide one possible explanation for these findings^7,8^. Studies show that as CAD progresses, changes in the levels of inflammatory biomarkers such as C-Reactive protein (CRP), leukocytes, monocytes and inflammation associated prothrombotic markers including platelets, can be observed^9–11^. Similarly, onset and severity of depression is associated with changes in the levels of immune and inflammatory factors including leukocytes, CRP, and platelets^12–22^. Neuroinflammation and peripheral inflammation are hypothesized to play an important role in both MDD and CAD respectively, providing a potential common biological pathway that may link neuroinflammation in depression together with atherosclerotic inflammation in CAD^7,8^.

Consortium efforts over the past decade have yielded results from large genome-wide association studies of depression, and independently, CAD^23,24^. These studies provide a map of statistical associations between common single nucleotide polymorphisms (SNPs) and each diagnosis, respectively. However, the genes that are implicated by these SNP associations must also be identified to define the biological pathways that might be shared between these conditions. Transcriptome wide association scans (TWAS) employ functional data to map SNPs to genes based on their ability to regulate gene expression^25^. In this study, we first used the TWAS method S-MultiXcan to map depression and CAD SNP-associations to the genes they regulate using cross-tissue expression quantitative trait loci (eQTL) annotations^25^. We then performed a series of enrichment and pathway analyses to characterize the gene set that was significantly associated with both depression and CAD. Our findings suggest that genes associated with both CAD and depression are enriched for both inflammatory pathways and genes previously associated with cardiomyopathy.

Next, we developed a clinical hypothesis based on these findings and tested it in three separate large electronic health records (EHR) data sets –Vanderbilt University Medical Center EHR, All of Us Research Program, and Mass-General Brigham EHR. At each site, we find that the rate of prevalent cardiomyopathy is significantly higher in subjects with (m)dCAD compared to those with CAD alone. Taken together, results of our study link depression and CAD through genetically controlled inflammatory mechanisms, and further suggest a relationship between (m)dCAD and cardiomyopathy.

## Materials and Methods

### Genome-wide Association Study Data – Coronary Artery Disease

We utilized publicly available summary statistics from a recently published GWAS of CAD as the foundation for our MetaXcan and S-MultiXcan analyses^23^. This study included a genome-wide meta-analysis of CAD cases and controls from the UK Biobank (UKBB) and CARDIoGRAMplusC4D^23^. CAD was defined in the UKBB by ICD10 codes (I21 – I25), procedural codes (K40-K46, K49, K50 and K75), and self-report of heart attack/myocardial infarction, coronary angioplasty +/- stent, coronary artery bypass graft surgery, or triple heart bypass. Controls were defined by the absence of features used to define cases and the absence of any family history of “heart disease”. Application of these criteria resulted in 34,541 CAD cases and 261,984 controls. The second sample used in this meta-analysis was the CARDIoGRAMplusC4D GWAS which itself was a meta-analysis of twenty-eight studies of European or South Asian descent populations. CAD was operationally defined differently across the cohorts, but all definitions relied on clinical or procedural markers of CAD, evidence of myocardial infarction, or imaging data consistent with a CAD diagnosis (<u>PMC3679547).</u> The CARDIoGRAMplusC4D GWAS included 63,746 CAD cases and 130,681 controls. The meta-analysis of UKBB and CARDIoGRAMplusC4D totaled 122,733 CAD cases and 424,528 controls.

### Genome-wide Association Study Data – Depression

We used summary statistics from the PGC and UKBB as the base for the MetaXcan analysis of depression^24^. The genome-wide association meta-analysis for depression included 170,756 cases and 329,443 controls. These results included summary statistics from a meta-analysis of the 33 cohorts of the Psychiatric Genomics Consortium as described in Wray et al. (2018), (https://doi.org/10.1038/s41588-018-0090-3) and the broad depression phenotype in the full release of the UKBB as described in (Howard et al. (2018), https://doi.org/10.1038/s41467-018-03819-3). The broad depression phenotype in UKBB included self-reported help-seeking behavior for “nerves, anxiety, tension or depression (once at any visit)” from a general practice physician or a psychiatrist. Secondly, a subject was defined as a case if there was a primary or secondary diagnosis of a depressive mood disorder from linked hospital admission records (UK Biobank fields: 41202 and 41204; ICD codes: F32—Single episode depression, F33—Recurrent depression, F34—Persistent mood disorders, F38—Other mood disorders and F39—Unspecified mood disorders). The remaining respondents were classed as controls if they answered “No” to both questions on all assessments. For the PGC sample, cases were those who met international consensus criteria (DSM-IV, ICD-9, or ICD-10) for a lifetime diagnosis of MDD established using structured diagnostic instruments from assessments by trained interviewers, clinician-administered checklists, or medical record review. Controls in most samples were screened for the absence of lifetime MDD (22/29 samples) or randomly selected from the population.

### Statistical Analysis of Genetic Data

#### S-MultiXcan Analysis of Depression and CAD GWAS Summary Statistics

Predicted expression models of 22,207 genes across 49 tissues were developed (supplementary materials) as previously described using data from the Genotype Tissue Expression Project^25,26^. These models were then used to train gene-based predicted associations for each set of summary statistics described above^25,26^. The subsequent associations with CAD and depression were tested using a multivariate regression model (Summary-MultiXcan or S-MultiXcan) which uses cross-tissue eQTL information^25^. S-MultiXcan infers the gene-level MultiXcan association results, using univariate S-PrediXcan results and LD information from a reference panel (i.e., the GTEx data set). Correlations between tissues are accounted for using a pseudo-inverse approach which applies a singular value decomposition (SVD) of the covariance matrix to keep only the components of large variation.

We used a Bonferroni correction to adjust the statistical significance threshold for 20,971 gene-based tests in the CAD MetaXcan analysis (p < 2.38e-06) and 20,945 gene-based tests in the depression MetaXcan analysis (p < 2.38e-06).

#### Enrichment Analysis of Gene-based Association Results (Depression and CAD)

After identifying genes associated with either depression or CAD, we determined whether genes associated with both CAD and depression (n=185) were more abundant than expected by chance, based on the number of genes significantly associated with each phenotype independently. We performed a hypergeometric test using the hyper R package and the publicly available website http://nemates.org/MA/progs/overlap_stats.html.

#### Gene Set Enrichment Analyses (GSEA)

We used the Gene Set Enrichment Analysis Web-based Tool (http://www.gsea-msigdb.org/gsea/msigdb/annotate.jsp) to functionally annotate the pathways for the 185 genes associated with *both* depression and CAD. We restricted our analysis to the ‘canonical pathways’ class with FDR q-values less than 0.05 and the GTEx compendium expression profiles. There were 2,982 pathways, composed of 40,786 genes, tested in the null ‘background’ set, provided by GSEA.

### Description of Clinical Populations

#### Vanderbilt University Medical Center (VUMC) Electronic Health Record (EHR)

Vanderbilt University Medical Center (VUMC) is a tertiary care center that provides inpatient and outpatient care in middle Tennessee and surrounding communities. The VUMC electronic health record (EHR) system was established in 1990 and includes data on billing codes from the International Classification of Diseases, 9th and 10th editions (ICD-9 and ICD-10), Current Procedural Terminology (CPT) codes, laboratory values, reports, and clinical documentation. A fully de-identified mirror image of the EHR, called the Synthetic Derivative, is available to Vanderbilt faculty for research purposes^27^.

#### Massachusetts General Brigham (MGB) EHR

Mass General Brigham (MGB) is a hospital network that includes Massachusetts General Hospital, Brigham and Women’s hospital, and other community and specialty hospitals in Boston area. The data source of MGB EHR is MGB Research Patient Data Registry (RPDR; https://rpdrssl.partners.org), an EHR database which spans more than 20 years of data from over 6.5 million patients and includes data on diagnoses (billing codes; ICD-9 and ICD-10 codes), procedures (CPT codes), laboratory values, and clinical notes. Data floors were applied to the MGB RPDR to include only patients with at least one clinical note and three visits since 2005 with visit more than 30 days apart.

#### All of Us Research Program

We used data from the *All of Us* Research Program, a multi-ancestry population-based cohort that contains various forms of information on individuals^28^. This data repository enrolls participants 18 years of age or older and contains their demographic and medical record-based information. The data for disease diagnosis was collected using billing codes and was the converted to phecodes.

#### Phenotypic analysis in the Synthetic Derivative

Informed by results of the genomic analyses, we first investigated the prevalence of cardiomyopathy in three mutually exclusive groups of patients in the VUMC-EHR including those who had a diagnosis of (1) depression or major depression *without* comorbid CAD (dep), (2) CAD *without* comorbid depression (CAD), and (3) CAD *with* comorbid depression or major depression ((m)dCAD)(Table S1). To reduce missingness of clinical data and enrich the sample for patients who receive their primary care at VUMC, we applied a simple “medical home” heuristic requiring the presence of any five codes on different days over a period of at least three years. We selected the cohort of individuals meeting these criteria who also had not been included in the VUMC BioBank which resulted in a total of 988,002 individuals. The genotyped sample was held out of this analysis to maintain its independence for future related research studies including genomic data.

We defined dep-only, CAD-only, and (m)dCAD using phecodes which are higher order combinations of at least two related ICD codes, occurring on two different days, using the R PheWAS package. Specifically, “CAD-only” cases required the presence of phecode 411.4 and absence of 296.2 phecode (N = 47,351), “dep-only” cases included phecode 296.2 and absence of 411.4 phecode (N = 57,069) and (m)dCAD required the presence of a phecode for both CAD and depression (N = 6,725). We next calculated the prevalence of cardiomyopathy, indicated by the presence of one or more of the following phecodes “425.1”, “425.11”, “425.12”, “425.8”, “425”. We then compared the prevalence of cardiomyopathy diagnoses between each diagnostic group using Pearson’s Chi-squared test and Fisher’s exact T-test.

We then used a logistic regression model to determine whether cardiomyopathy was indeed more prevalent among patients with (m)dCAD, compared to CAD alone, after controlling for potential confounding. Cases were defined as those with cardiomyopathy, controls were those without cardiomyopathy. The exposure was defined as depression, and features known to associate with both depression and cardiomyopathy, age, race, sex, record median BMI, type 2 diabetes (T2D; Phecode = 250.2), tobacco use disorder (TUD; Phecode = 318) and hypertension (Phecodes = 401, 415.21, 453), were included as covariates.

Next, we used the same regression-based approach to test whether *incident* cardiomyopathy was increased among individuals with (m)dCAD compared to CAD alone. All individuals with cardiomyopathy coded prior to depression or CAD (N = 14,583) were excluded. Hence the cardiomyopathy exposure was required to be coded after both depression and CAD. This resulted in a total of 2,345 cardiomyopathy cases and 45,330 cardiomyopathy controls. We then fitted two multivariable logistic regression models to test whether depression increased the odds of incident cardiomyopathy among patients with (m)dCAD compared to patients with CAD alone, after adjusting for the same previously described confounders.

### Sensitivity analyses using Severe Depression

Previous literature points towards progression and worsening of CAD with increasing depression severity. To test whether cardiomyopathy more commonly co-occurred with severe depression, we redefined our exposure using a stricter MDD definition requiring the presence of 296.22 phecode and ICD9/10 codes (Table S8) which dropped the sample size from 57,069 depression-exposed patients to 41,423 MDD-exposed patients. The same regression models described above were fitted to the data.

### Phenotypic Replication in MGB and *All of Us* Data

We replicated the EHR analyses in two external cohorts, *All of Us* and MGB. We used the same phecode based definitions to define our inclusion and exclusion criteria, case and control labels, and exposures. Sensitivity analyses were similarly performed in these data by restricting the exposure definition to include individuals with only severe depression.

## Results

### Genes Associated with Both Depression and CAD

We intersected the 22,207 predicted expression gene models with the summary statistics of CAD and depression phenotypes as described above. While the tissues that best predicted gene expression differed between CAD and depression (Figure S4); the mean (Spearman’s rank = 0.433), standard deviation (Spearman’s rank = 0.565), minimum (Spearman’s rank = 0.389), and maximum (Spearman’s rank = 0.373) association Z-score statistics demonstrated strong correlation between depression and CAD across all tissues tested (Figure S3).

Results from the 20,971 gene-based tests of association with CAD, and 20,945 gene-based tests of association with depression are available in Table S2 and Table S3, respectively^23,24^. We identified a total of 1,455 genes significantly associated with CAD (p < 2.38e-06) and 928 genes significantly associated with depression (p < 2.38e-06) illustrated in Figures S1 and S2, respectively. We next identified the genes that were significantly associated with *both* depression and CAD. A total of 20,944 genes were tested in common between depression and CAD, and of these, 185 genes identified by their gene names were significantly associated with *both* CAD and depression (Figure 2; Table S4). The genes associated with both depression and CAD were distributed across the genome (Figure 2).

**Figure 1.**
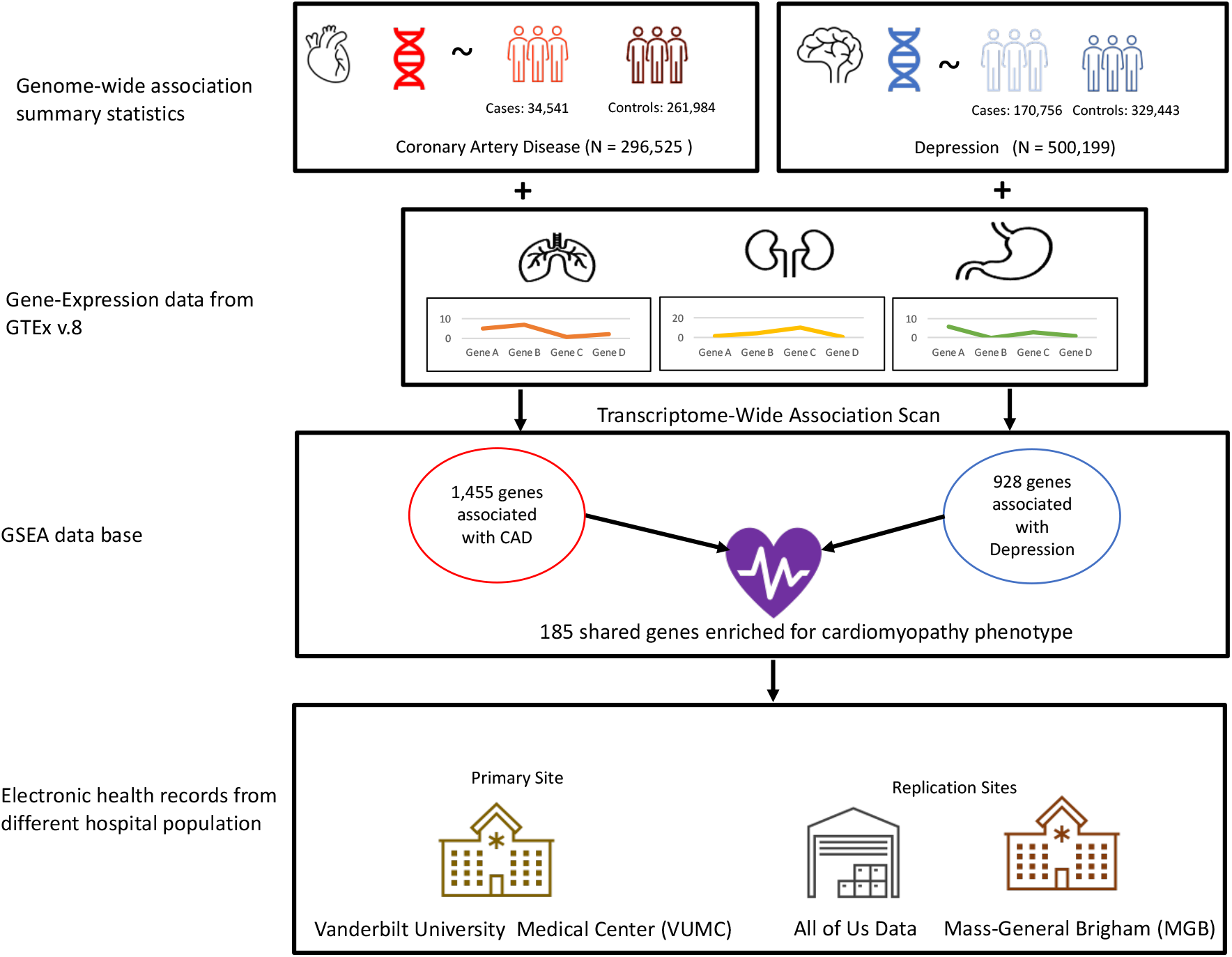
Overall Schematic of the Study. We performed a transcriptome-wide association study (TWAS) of genetically regulated expression (GReX) using the GTEx v.8 database and genome-wide summary statistics of Depression and Coronary Artery Disease separately. We derived genes that were shared between Depression and CAD, we performed pathway and enrichment analyses. Our results show that genes associated with both depression and CAD are associated with cardiomyopathy associated phenotypes.

**Figure 2.**
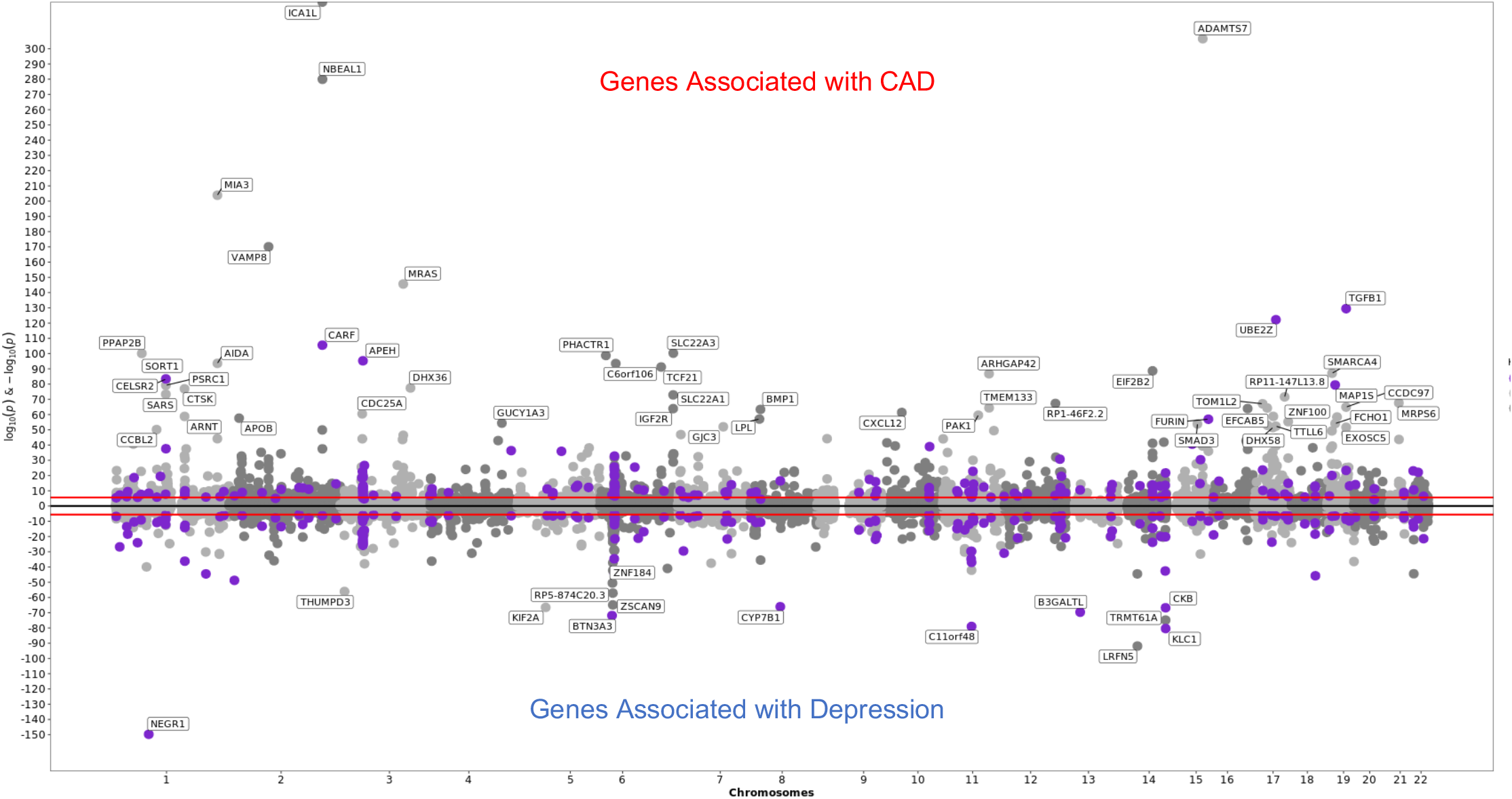
Genes associated with Depression and CAD using S-MultiXcan. A Miami plot of genes associated with both Depression and CAD via S-MultiXcan Each data point represents a gene grouped by chromosome (x-axis) and p value (y-axis) of the association with the phenotype. The top half of the graph are genes associated with CAD and the lower half of the graph represents genes associated with Depression. The purple dots represent the 185 genes shared between Depression and CAD. Genes with –log(p values) greater that 50 are labelled. The light and dark grey represents the non-shared genes on alternating chromosomes with light grey with being the non-shared genes on odd chromosomes and dark grey representing the non-shared genes on the even numbered chromosomes

### Enrichment and Pathway Analysis of 185 Genes Associated with Both Depression and CAD

We found that there were three times more genes associated with both CAD and depression than expected by chance given the number of genes significantly associated with each phenotype independently (hypergeometric test, p<1.718e-43) (Figure 3). Inflammatory (FDR q-value < 0.05) and cardiomyopathy-associated pathways (FDR q value < 0.05) were overrepresented among genes associated with both CAD and depression (Table 1). Five genes from the shared set of 185 (2.7%) were annotated to cardiomyopathy-associated pathways. Four of the 5 genes (*ATP2A2, ITGB4, SGCD*, and *CACNB4)* were annotated to all three cardiomyopathy pathways (Table S4). Thirteen additional genes (7%) were annotated to adaptive immune pathways (Table S4).

**Figure 3.**
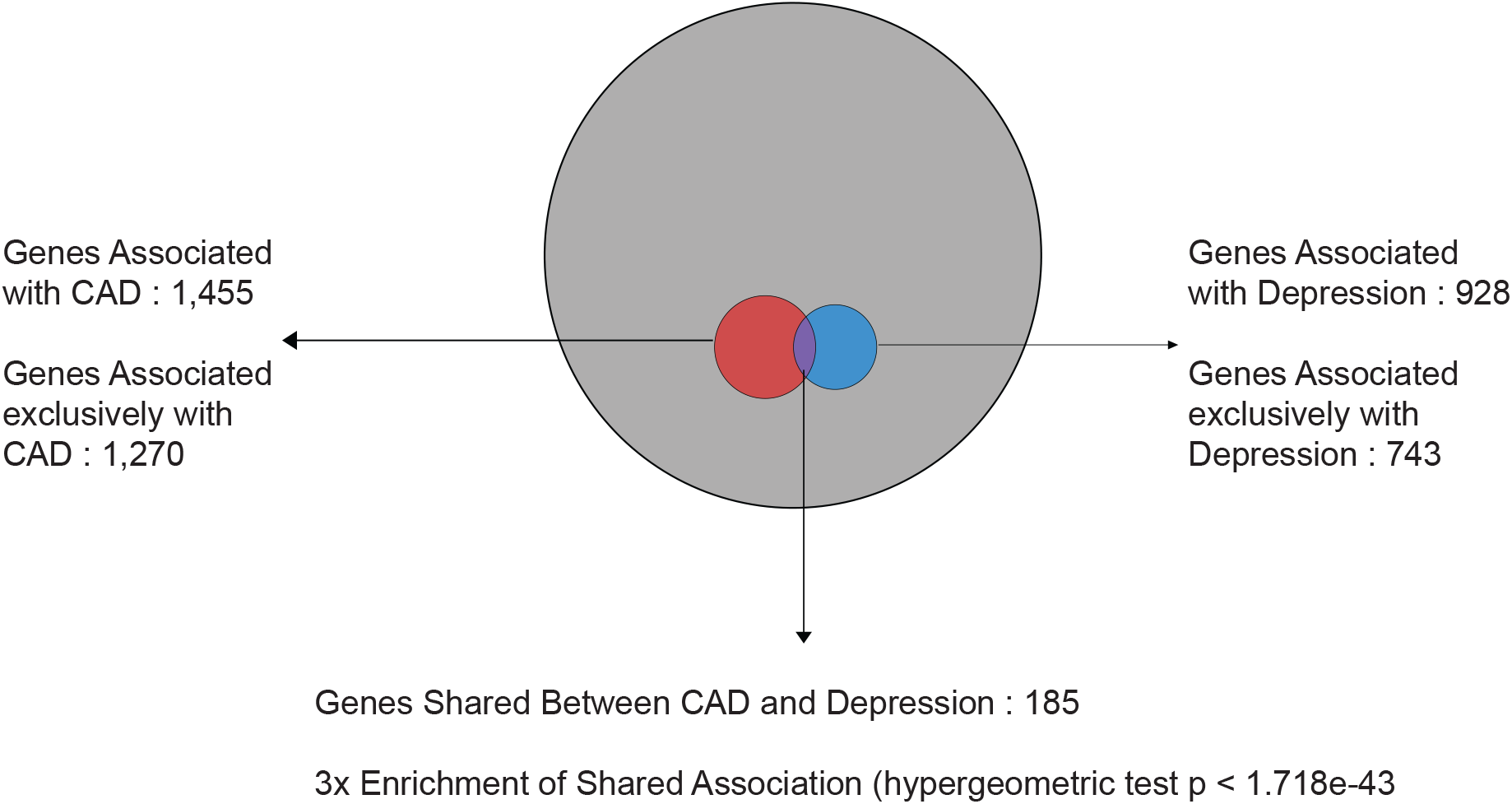
Enrichment test of the genes shared between Depression and CAD. Eulerr Plot to highlight the results of enrichment test of the genes associated with both depression and CAD. The grey circle represents the set of 20,944 genes that were tested for association with both phenotypes. The red portion represents the 1,270 genes significantly associated with CAD only. The blue portion represents the 743 genes significantly associated with depression only. The purple portion represents the 185 genes associated with both depression and CAD. Red + purple represents all CAD associated genes (1455) and blue + purple represents all genes associated with depression (928).

**Table 1:**
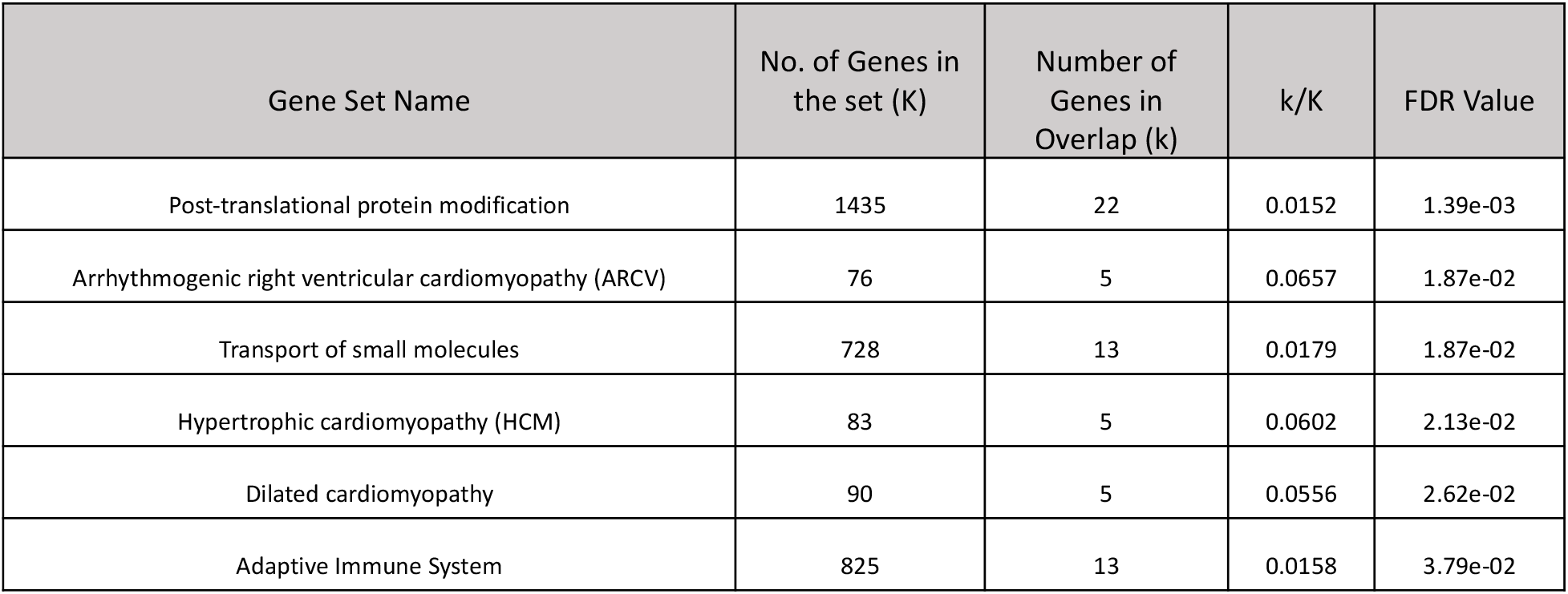
Gene Set Enrichment Analysis of the Shared Genes We used the 185 genes shared between Depression and CAD as the input for GESA (Canonical Pathways) browser and identified that the shared genes are associated with immune and cardiomyopathy associated phenotypes. K refers to the number of genes in the GSEA database for that particular pathway. k refers to the number of genes from our shared 185 gene-set in the particular pathway

| Gene Set Name | No. of Genes in the set (K) | Number of Genes in Overlap (k) | k/K | FDR Value |
| --- | --- | --- | --- | --- |
| Post-translational protein modification | 1435 | 22 | 0.0152 | 1.39e-03 |
| Arrhythmogenic right ventricular cardiomyopathy (ARCV) | 76 | 5 | 0.0657 | 1.87e-02 |
| Transport of small molecules | 728 | 13 | 0.0179 | 1.87e-02 |
| Hypertrophic cardiomyopathy (HCM) | 83 | 5 | 0.0602 | 2.13e-02 |
| Dilated cardiomyopathy | 90 | 5 | 0.0556 | 2.62e-02 |
| Adaptive Immune System | 825 | 13 | 0.0158 | 3.79e-02 |

**Table 2:** Demographic table for Phenotypes in VUMC This table describes the demographics for Depression, Major Depression, CAD, Comorbid Depression-CAD and Comorbid Major Depression-CAD in VUMC EHR data.

|  | Depression (broad) | Major Depression | CAD | Depression and CAD | Major Depression and CAD |
| --- | --- | --- | --- | --- | --- |
| *Sample sizes* |  |  |  |  |  |
| Total (N) | 57,069 | 41,423 | 47,349 | 6,725 | 4,175 |
| Cardiomyopathy | 809 (1.41%) | 537 (1.29%) | 5,314 (11.22%) | 1,001 (14.88%) | 589 (14.10%) |
| *Demographics* |  |  |  |  |  |
| Female (%) | 68.64 | 69.1 | 32.86% | 48.91% | 47.04% |
| Median Age, years (IQR) | 41(24.5 – 65.5) | 39 (22.5 – 55.5) | 68 (60 – 76) | 65 (56.5-72.5) | 64 (56 – 72) |
| Median BMI (IQR) | 27.34 | 27.28 | 28.81 | 29.29 | 29.68 |
| Underweight | 2,071 (3.63%) | 1,655 (4.0%) | 377 (0.79%) | 64 (0.95%) | 39 (0.93%) |
| Normal | 17,437 (30.56%) | 12,964 (31.3%) | 8,673 (18.31%) | 1,320 (19.62%) | 794 (19.02%) |
| Overweight | 14,154 (24.80%) | 10,204 (24.63%) | 15,520 (32.77%) | 1,979 (29.43%) | 1,291 (30.93%) |
| Obese | 19,247 (33.73%) | 14,203 (34.28%) | 17,423 (36.8%) | 2,812 (41.81%) | 1,951 (46.73%) |
| Unknown | 4,160 (7.29%) | 2,397 (5.78%) | 5,356 (11.31%) | 550 (8.18%) | 100 (2.4%) |
| *Cardiovascular Comorbidities* |  |  |  |  |  |
| Hypertension (N, %) | 19,784 (34.66%) | 13,513 (32.62%) | 38,905 (82.17%) | 6,200 (92.2%) | 3,837 (91.9%) |
| Type 2 Diabetes (N, %) | 7,452 (13.06%) | 5,116 (12.35%) | 15,207 (32.11%) | 3,227 (47.99%) | 2,062 (49.39%) |
| Tobacco Use Disorder (N, %) | 8,363 (14.65%) | 6,555 (15.82%) | 8,482 (17.91%) | 2,399 (35.67%) | 1,816 (43.5%) |

### Clinical Cardiomyopathy in (m)dCAD

Analyses performed in the EHR returned significant differences in the prevalence of cardiomyopathy between depression-only, CAD-only and (m)dCAD cohorts (Chisq P < 2.2e-16) (Table S5). Almost 15% of individuals with (m)dCAD had a cardiomyopathy diagnosis, compared to 11.2% in the CAD cohort and only 1.59 % in the depression cohort. The results remained significant when we compared only the (m)dCAD group to the CAD group (Fisher’s exact test p < 2.2e-16) (Table S5). We then replicated these findings in the MGH EHR and the All of Us Data. We next restricted the depression phenotype to major depression (phecode 296.22) and observed the same pattern in the prevalence of cardiomyopathy between the groups (Table S6).

### Prevalence of Clinical Cardiomyopathy after Adjusting for known Confounders

Results of the multivariable logistic model indicate that there is a 27% increase in the odds of prevalent cardiomyopathy with a diagnosis of (m)dCAD compared to CAD after adjusting for age, race, sex, hypertension diagnosis, smoking status, type-2-diabetes and BMI (see Methods for details) (p-value = 3.6e-09, OR = 1.27, 95% CI = 1.17-1.3). This finding was replicated in the *All of Us* (p-value = 0.012, OR = 1.16, 95%CI = 1.03-1.30) and MGH (p-value < 2e-16, OR = 1.32, 95%CI = 1.28-1.37) data sets.

Sensitivity analyses restricting the criteria for depression to “major depression” preserved the effect estimate and significance of the original finding (OR = 1.14, 95%CI = 1.06-1.26, p = 6.5e-03)(Figure 4). This finding was again replicated in the MGH cohort (OR = 1.23, 95%CI = 1.13-1.18, p < 2e-16). While the effect estimate was similar in the *All of Us* data set, the finding did not reach statistical significance likely due to a reduced sample size (OR = 1.13, 95%CI = 0.97-1.32, p-value = 0.12)

**Figure 4.**
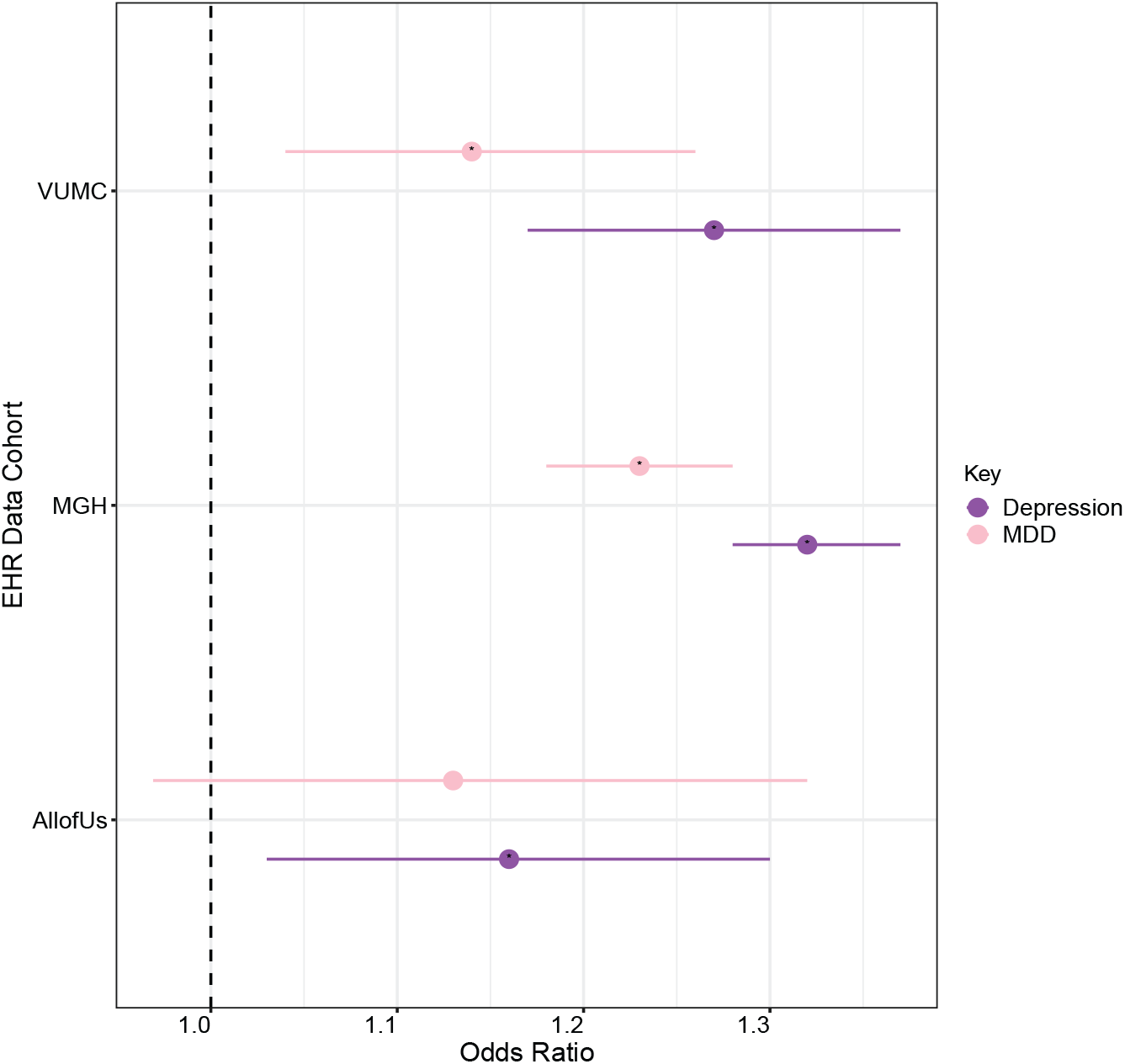
Forest plot illustrating odds of prevalent cardiomyopathy in (m)dCAD compared to CAD. The association between (m)dCAD vs CAD and prevalent cardiomyopathy after controlling for race, median age across the medical record, sex, T2D, hypertension, and smoking (ever/never) at each of the three sites (VUMC, MGB, and All of Us). Purple represents results from an analysis in which depression was broadly defined while light pink represents results from an analysis restricted to major depression. Whiskers indicate 95% CIs.

### Rate of Incident Cardiomyopathy in (m)dCAD compared to CAD

Lastly, we tested the hypothesis that (m)dCAD is a risk factor for the development of *subsequent* cardiomyopathy. Using a multivariable logistic regression model, we found no increase in the odds of incident cardiomyopathy among (m)dCAD patients compared to CAD patients, after adjusting for known confounders in the VUMC EHR (OR = 0.96, 95% CI = 0.84-1.09, p-value = 0.53) (Figure 5) and in the *All of Us* data (OR = 0.864, 95%C.I. = 0.730-1.021, p-value = 0.086). In the MGH data, not only did we observe a lack of evidence for increased incident cardiomyopathy subsequent to (m)dCAD, in fact we observed that (m)dCAD cases were significantly *less* likely to be diagnosed with new onset cardiomyopathy compared to CAD alone (OR = 0.783, 95%C.I. = 0.743-0.825, p-value < 2e-16).

**Figure 5.**
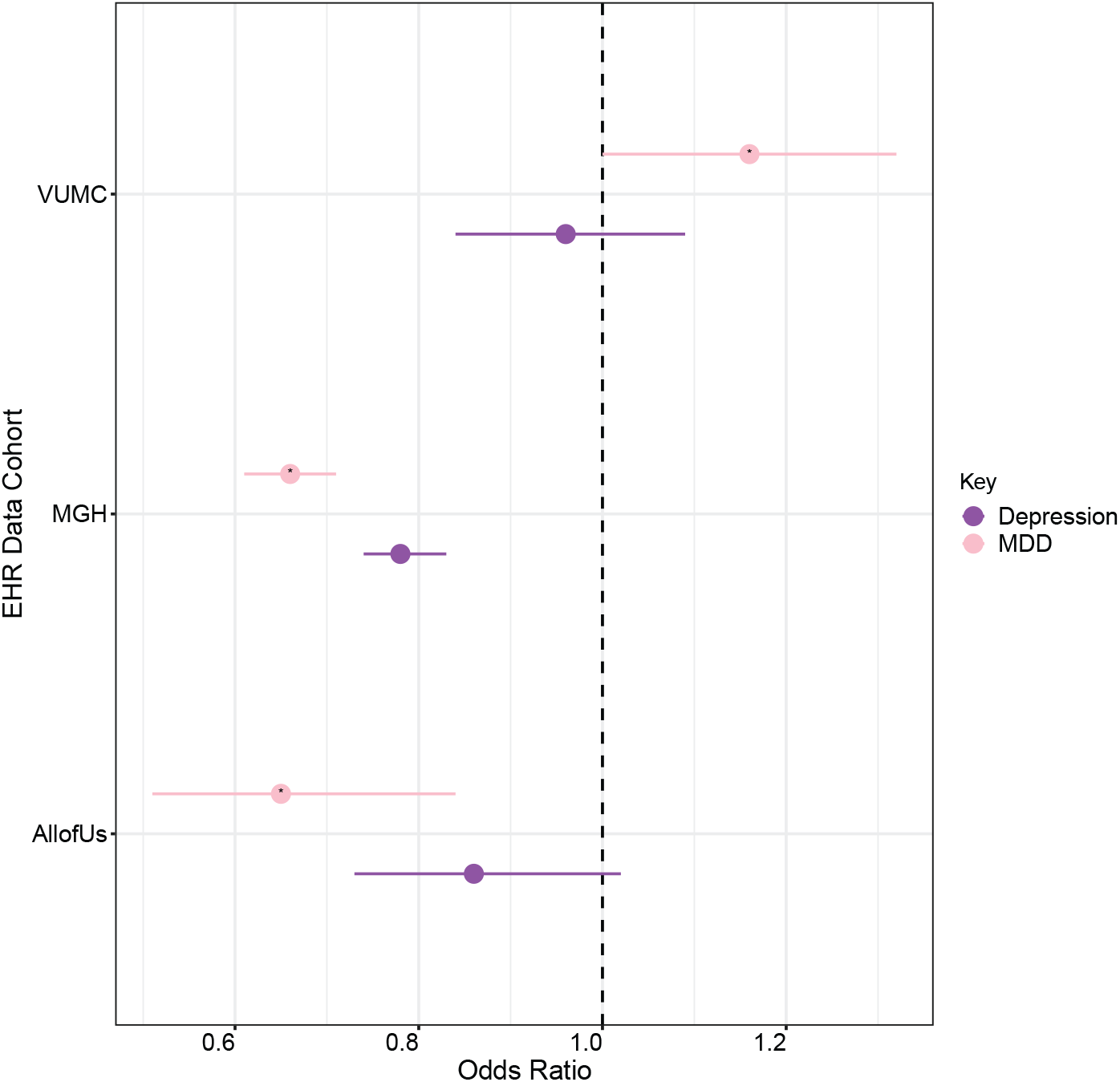
Forest plot illustrating odds of incident cardiomyopathy in (m)dCAD compared to CAD. The association between (m)dCAD vs CAD and incident cardiomyopathy after controlling for race, median age across the medical record, sex, T2D, hypertension, and smoking (ever/never) at each of the three sites (VUMC, MGB, and All of Us). Purple represents results from an analysis in which depression was broadly defined while light pink

After restricting the definition of depression to “major depression” in the VUMC data, the odds of incident cardiomyopathy were elevated but not significantly so (OR = 1.16, 95% C.I. = 1.00-1.34, p-value = 0.04). However, again, in the *All of Us* (OR = 0.66, 95% C.I. = 0.51-0.84, p-value = 9e-4) and MGH (OR = 0.54, 95% C.I. = 0.50-0.58, p-value < 2 e-16) data, the rate of incident cardiomyopathy was lower in (m)dCAD than in CAD alone.

## Discussion

Depression and CAD are highly comorbid conditions and the molecular mechanisms underlying this common comorbidity remain understudied. Here we present an integrative genotypic approach to identify genes that map to SNP associations for each condition, followed by a phenotypic analysis to test a hypothesis motivated by the genetic findings. By performing a cross-tissue transcriptome-wide association scan using large scale GWAS data for depression and CAD, we identified 185 genes (three time more than expected by chance) that were significantly associated with both depression and CAD. Interestingly, the genes associated with both phenotypes were not necessarily those most strongly associated with depression or CAD independently. On balance, these results suggest that comorbid (m)dCAD may have a partially unique genetic architecture which overlaps a subset of genes associated with depression and CAD.

Pathway analyses indicate that the genes involved in (m)dCAD are enriched for inflammatory and cardiomyopathy-associated pathways. Specifically, we observed an enrichment for adaptive immune system genes which can be understood in the context of previous studies that have highlighted immune dysfunction in both depression and CAD^15,29–36^. Moreover, research has shown that depression is associated with low-grade chronic inflammation whereas CAD has phases of both, acute and chronic inflammation^36–38^. Hence the enrichment of adaptive immune system genes suggests that depression and CAD could be linked through exposure to low-grade chronic inflammation.

There were six genes *ATP2A2, ITGB4, SGCD, CACNB4, ACTN2*, and *TGFB1* associated with both CAD and depression, which are known to be involved in cardiomyopathy (Table S4). Differential expression of *ATP2A*, which encodes a Ca^2+^ pump on the endoplasmic reticulum, is associated with cardiac phenotypes including dilated cardiomyopathy^39,,40^; and heterozygous conditional knockout mice demonstrate an essential role for *ATP2A* in neuronal calcium homeostasis resulting in behavioral phenotypes^41^. Similarly, *CACNB4* is the one of the most abundant voltage-gated calcium channel subunits expressed in the brain where it is critical for presynaptic signaling, and co-located in the heart where early data suggests it may be involved in cardiac contraction^42–44^. The gene locus was also recently implicated in idiopathic cardiomyopathy in individuals of African-American ancestry^45^. *ACTN2*, a cardiac-specific structural protein, is classically involved in familial hypertrophic, dilated, and arrhythmogenic cardiomyopathy^46–48^ while *TGFB1* is a cytokine with a variety of cellular functions implicated in cardiac hypertrophy and hypertrophic cardiomyopathy along with congenital heart disease^49,50^. Mutations in delta-sarcoglycan (*SGCD)* result in severe neuromuscular disease in humans and knockout mice develop early progressive cardiomyopathy and dilated cardiomyopathy^51,52^. However, less is known about the impact of differences in expression of these genes (*ACTN2, TGFB1*, and *SGCD*) on depression.

Interpreted in the context of previous studies, our genetic findings suggest that depression and CAD could share low grade chronic inflammation^36,53–55^. Moreover, genetic results of our study raised the hypothesis that a predisposition to both depression and CAD (clinically observed as (m)dCAD) may further predispose individuals to cardiomyopathy. Canonically, viral infections are considered the most common trigger and cause of inflammatory cardiomyopathy thus leading to immune mechanisms which potentially damage the myocardial function^56–63^. However, an alternative model consistent with our findings could position chronic low-grade inflammation as a shared risk factor for depression, CAD, and cardiomyopathy. We investigated the hypothesis that cardiomyopathy co-occurs more commonly with (m)dCAD than with CAD, by harnessing the power of large-scale electronic health record data at VUMC, MGB, and *All of Us*. These three systems represent both hospital ascertainment (VUMC and MGB) and volunteer ascertainment (*All of Us*). Results of these analyses suggest that while prevalent cardiomyopathy does in fact co-occur more frequently with (m)dCAD than with CAD alone, the order of events remains unclear. For example, it remains possible that (a) cardiomyopathy increases the risk of co-morbid depression in CAD, (b) treatment for depression provides some protection against subsequent cardiomyopathy, or (c) both cardiomyopathy and (m)dCAD share additional risk factors. While our work demonstrating the clinical co-occurrence suggests that cardiomyopathy patients may benefit from depression screening, future work is needed to disentangle their precise relationship and inform clinically translatable mitigation strategies.

Despite its strengths, our study is limited by the well-established caveats of EHR based phenotyping which itself suffers from individual bias of the clinician. TWAS analyses rely on SNP-based predictive models of mRNA trained using mostly European-ancestry individuals in GTEx v8 and assumes additivity of SNP effects on gene expression, which ignores the possibility of epistatic and gene-environment interactions. Online pathway tools are prone to knowledge bias in selection of the “validated” genes annotated to pathways.

Nevertheless, these analyses highlight the unique genetic and phenotypic architecture of (m)dCAD compared to CAD and depression alone. We add to the literature the observation that prevalent, but perhaps not incident cardiomyopathy, is also more common in patients with (m)dCAD compared to CAD alone. Importantly, this hypothesis was motivated by genetic findings suggesting that the genes associated with both depression and CAD are enriched for pathways with a known role in immune and cardiomyopathy-associated biological processes. This data-driven approach highlights the power of *in silico* transcriptome-wide studies to motivate hypotheses that can be tested in large EHR databases.

## Data Availability

The electronic health record data that support the findings of this study are available from Vanderbilt University Medical Center, but restrictions apply to the availability of these data, which were used under license for the current study, and so are not publicly available. Data are, however, available from the institution with appropriate material transfer agreements or data use agreements and permission of Vanderbilt University Medical Center.

## Non-Standard Abbreviations and Acronyms

CAD: Coronary artery disease
CI: Confidence interval
CAD: Coronary Artery Disease
CVD: Cardiovascular disease
HER: Electronic health record
GWAS: Genome-wide association study
ICD: International Classification of Diseases
LD: Linkage disequilibrium
MD: Major depression
MDD: Major depressive disorder
MHC: Major histocompatibility complex
MR: Mendelian randomisation
OR: Odds ratio
PGS: Polygenic score
SNP: Single nucleotide polymorphism
VUMC: Vanderbilt University Medical Center

## Funding

KS is funded by the American Heart Association Fellowship AHA827137. LKD is supported by R56MH120736. JWS and LKD are supported in part by NIMH R01 H118233. JMS is funded by 1F31MH124306-01A1. QSW was supported by NIH 1R01HL140074.

### CTSA (SD, Vanderbilt Resources)

The deidentified EHR used at VUMC was supported by the National Center for Research Resources, Grant UL1 RR024975-01, and is now at the National Center for Advancing Translational Sciences, Grant 2 UL1 TR000445-06. The content is solely the responsibility of the authors and does not necessarily represent the official views of the NIH.

### BioVU

The dataset(s) used for the analyses described were obtained from Vanderbilt University Medical Center’s BioVU which is supported by numerous sources: institutional funding, private agencies, and federal grants. These include the NIH funded Shared Instrumentation Grant S10RR025141; and CTSA grants UL1TR002243, UL1TR000445, and UL1RR024975. Genomic data are also supported by investigator-led projects that include U01HG004798, R01NS032830, RC2GM092618, P50GM115305, U01HG006378, U19HL065962, R01HD074711; and additional funding sources listed at https://victr.vumc.org/biovu-funding/.

The *All of Us* Research Program is supported by grants through the National Institutes of Health, Office of the Director: Regional Medical Centers: 1 OT2 OD026549; 1 OT2 OD026554; 1 OT2 OD026557; 1 OT2 OD026556; 1 OT2 OD026550; 1 OT2 OD026552; 1 OT2 OD026553; 1 OT2 OD026548; 1 OT2 OD026551; 1 OT2 OD026555; IAA#: AOD 16037; Federally Qualified Health Centers: HHSN 263201600085U; Data and Research Center: 5 U2C OD023196; Biobank: 1 U24 OD023121; The Participant Center: U24 OD023176; Participant Technology Systems Center: 1 U24 OD023163; Communications and Engagement: 3 OT2 OD023205; 3 OT2 OD023206; and Community Partners: 1 OT2 OD025277; 3 OT2 OD025315; 1 OT2 OD025337; 1 OT2 OD025276. In addition to the funded partners, the *All of Us Research Program* would not be possible without the contributions made by its participants.

## Declarations

### Contributions

K.S. and L.K.D. conceptualized and designed the work. K.S, J.M.S, and H.L. implemented the computational procedures and performed data analysis. N.J.C, Q.S.W., and E.C.H provided important clinical and intellectual insights. All authors read, edited, and approved the final manuscript.

## Ethics declarations

### Ethics approval and consent to participate

This study was reviewed by the VUMC IRB and designated as non-human subjects research because of the use of fully de-identified data (IRB# 190418 and IRB# 201609).

## Supplementary Materials

Supplementary Methods

Supplementary Figure 1

Supplementary Figure 2

Supplementary Figure 3

Supplementary Figure 4

Supplementary Table 1

Supplementary Table 2

Supplementary Table 3

Supplementary Table 4

Supplementary Table 5

Supplementary Table 6

## Supplementary Materials

### Description of Clinical Populations

#### Vanderbilt University Medical Center (VUMC) Electronic Health Record (EHR)

Vanderbilt University Medical Center (VUMC) is a tertiary care center that provides inpatient and outpatient care in middle Tennessee and surrounding communities. The VUMC electronic health record (EHR) system was established in 1990 and includes data on billing codes from the International Classification of Diseases, 9th and 10th editions (ICD-9 and ICD-10), Current Procedural Terminology (CPT) codes, laboratory values, reports, and clinical documentation. A fully de-identified mirror image of the EHR, called the Synthetic Derivative, is available to Vanderbilt faculty for research purposes. A data floor (i.e., “medical home”) heuristic of any five codes on different days over a period of at least three years was imposed to enrich the sample for primary care.

#### All of Us Research Program

All of Us Research Program, a population-based cohort that contains demographic, EHR, and survey information on participants. Participants enroll digitally through the All of Us website. After completion of the consent modules and enrollment, participants are given several health-measuring surveys to be completed. As of July 2019, the All of Us program had enrolled more than 175,000 core participants and more than 230,000 total participants.

**Figure S1.**
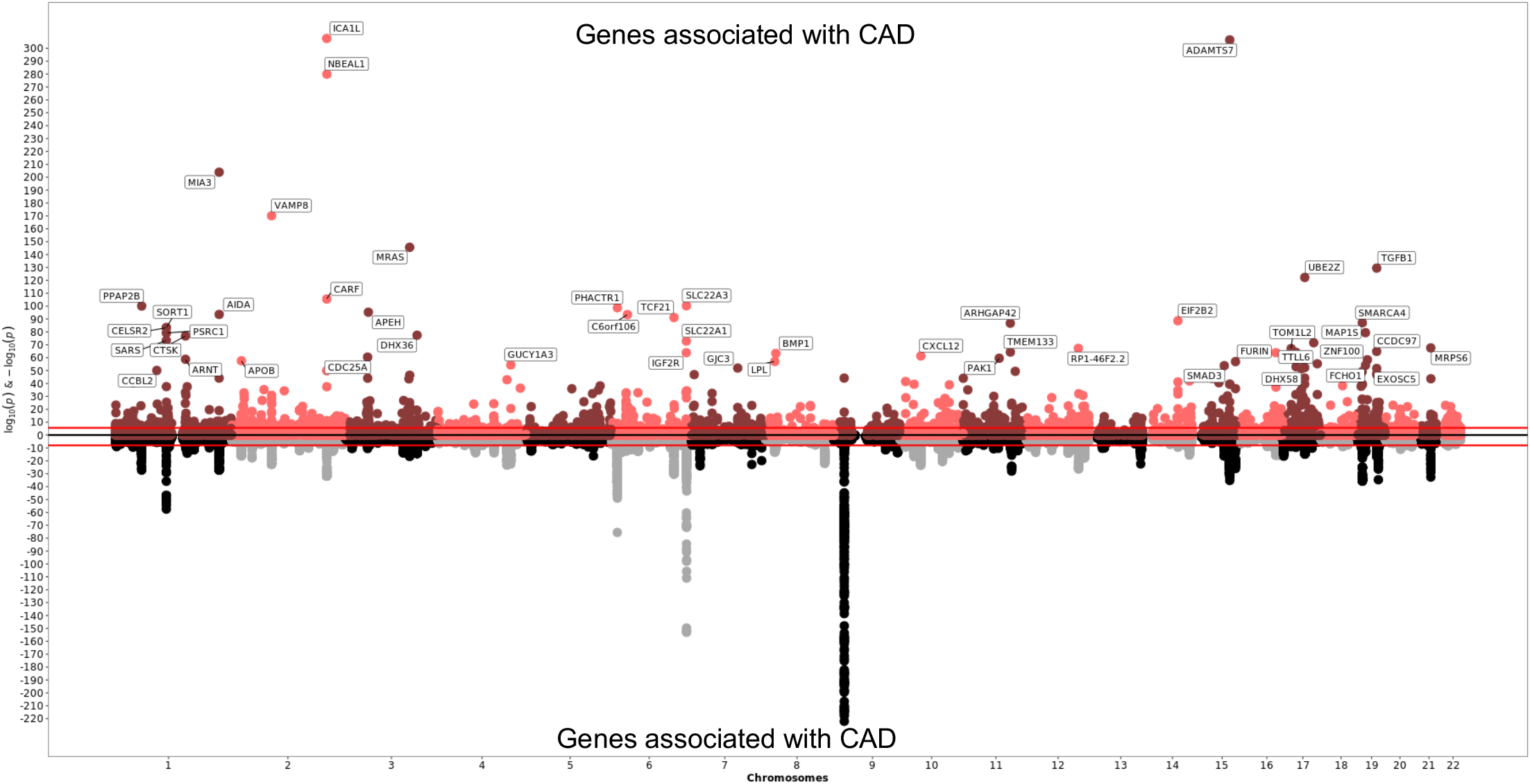
Genes associated with CAD using S-MultiXcan. A Miami plot of genes and SNPs associated with CAD via S-MultiXcan. Above the x-axis, each data point represents a gene grouped by chromosome (x-axis) and p value (y-axis) of the association with the phenotype. The light red and darl red are alternating colors for genes on odd and even chromosomes. Below the x-axis, each data point represents the SNPs associated with CAD from the input GWAS. Genes with –log(p values) greater that 50 are labelled. The black and grey are alternating colors for SNPs on odd and even chromosomes

**Figure S2.**
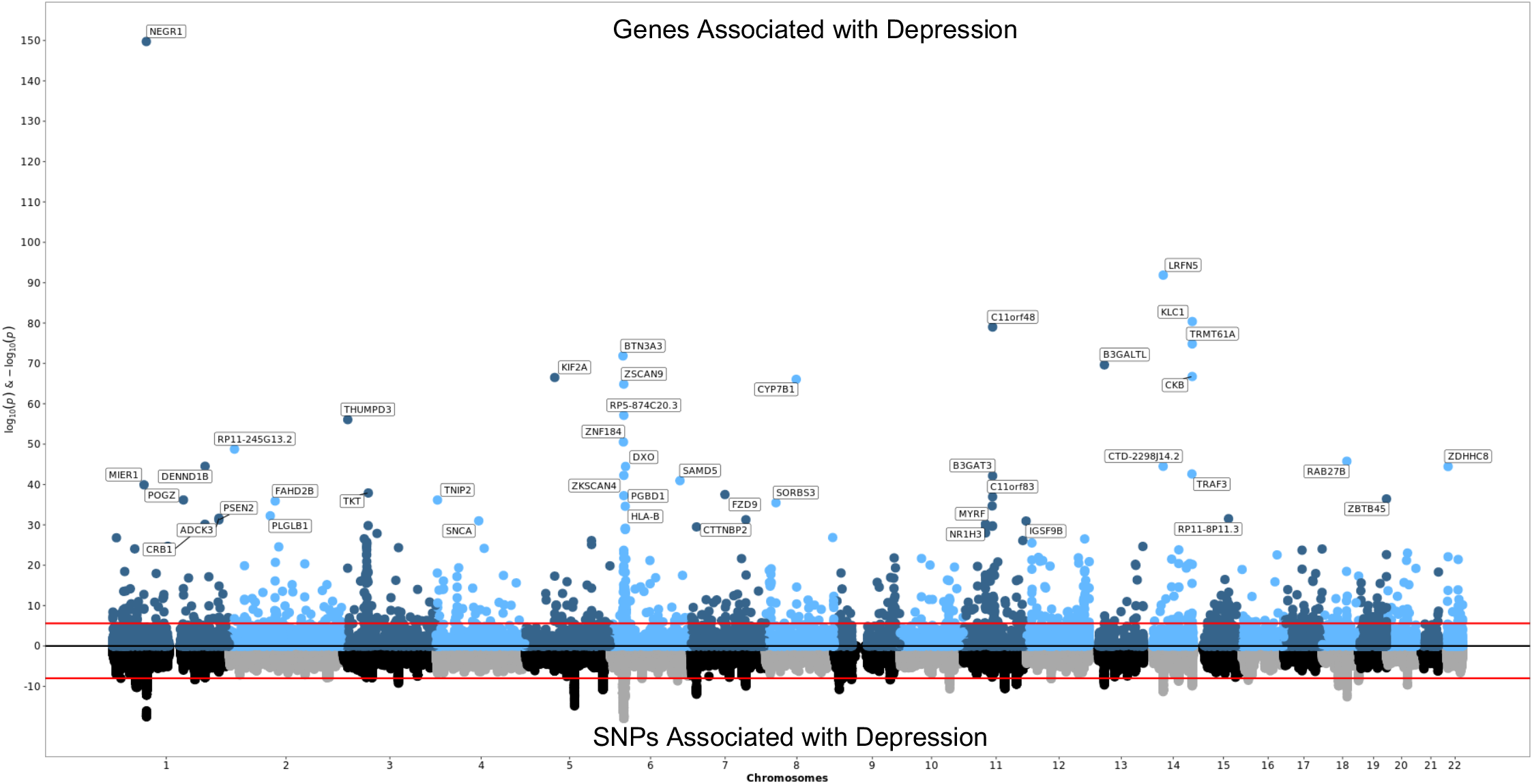
Genes associated with Depression using S-MultiXcan. A Miami plot of genes and SNPs associated with Depression via S-MultiXcan. Above the x-axis, each data point represents a gene grouped by chromosome (x-axis) and p value (y-axis) of the association with the phenotype. The light red and dark blue are alternating colors for genes on odd and even chromosomes Below the x-axis, each data point represents the SNPs associated with Depression from the input GWAS. Genes with –log(p values) greater that 50 are labelled. The black and grey are alternating colors for SNPs on odd and even chromosomes

**Figure S3.**
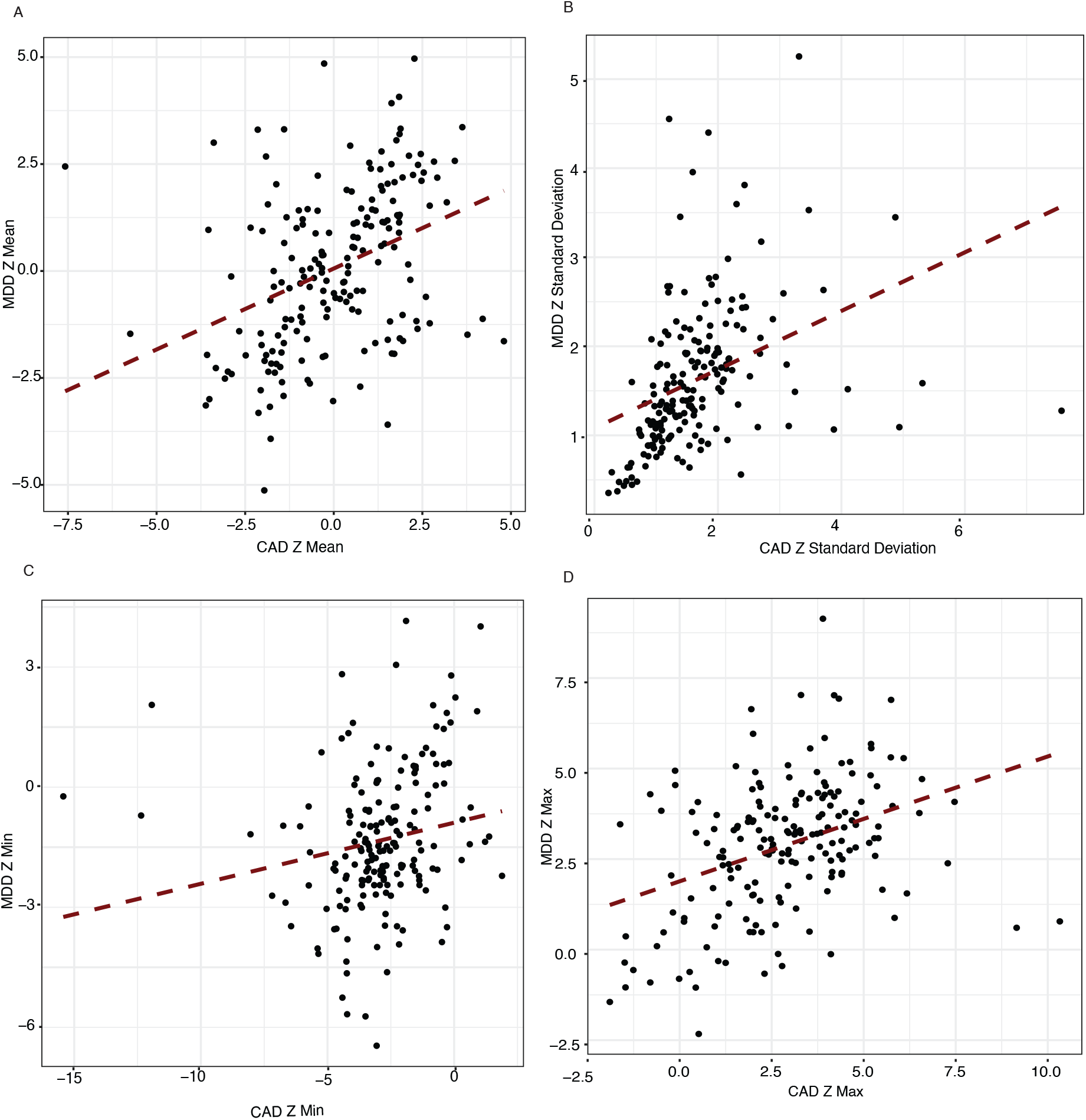
Correlation plots between the test statistics of Depression and CAD for the 185 genes associated with both the phenotypes. (a) Correlation Plot of Z Mean of Depression and CAD for the 185 genes shared between both the phenotypes (Spearman’s coefficient = 0.433) (b) Correlation Plot of Z Standard Deviation of Depression and CAD for the 185 genes shared between both the phenotypes (Spearman’s coefficient = 0.565) (c) Correlation Plot of Z Min of Depression and CAD for the 185 genes shared between both the phenotypes (Spearman’s coefficient = 0.389) (d) Correlation Plot of Z Max of Depression and CAD for the 185 genes shared between both the phenotypes (Spearman’s coefficient = 0.373)

**Figure S4.**
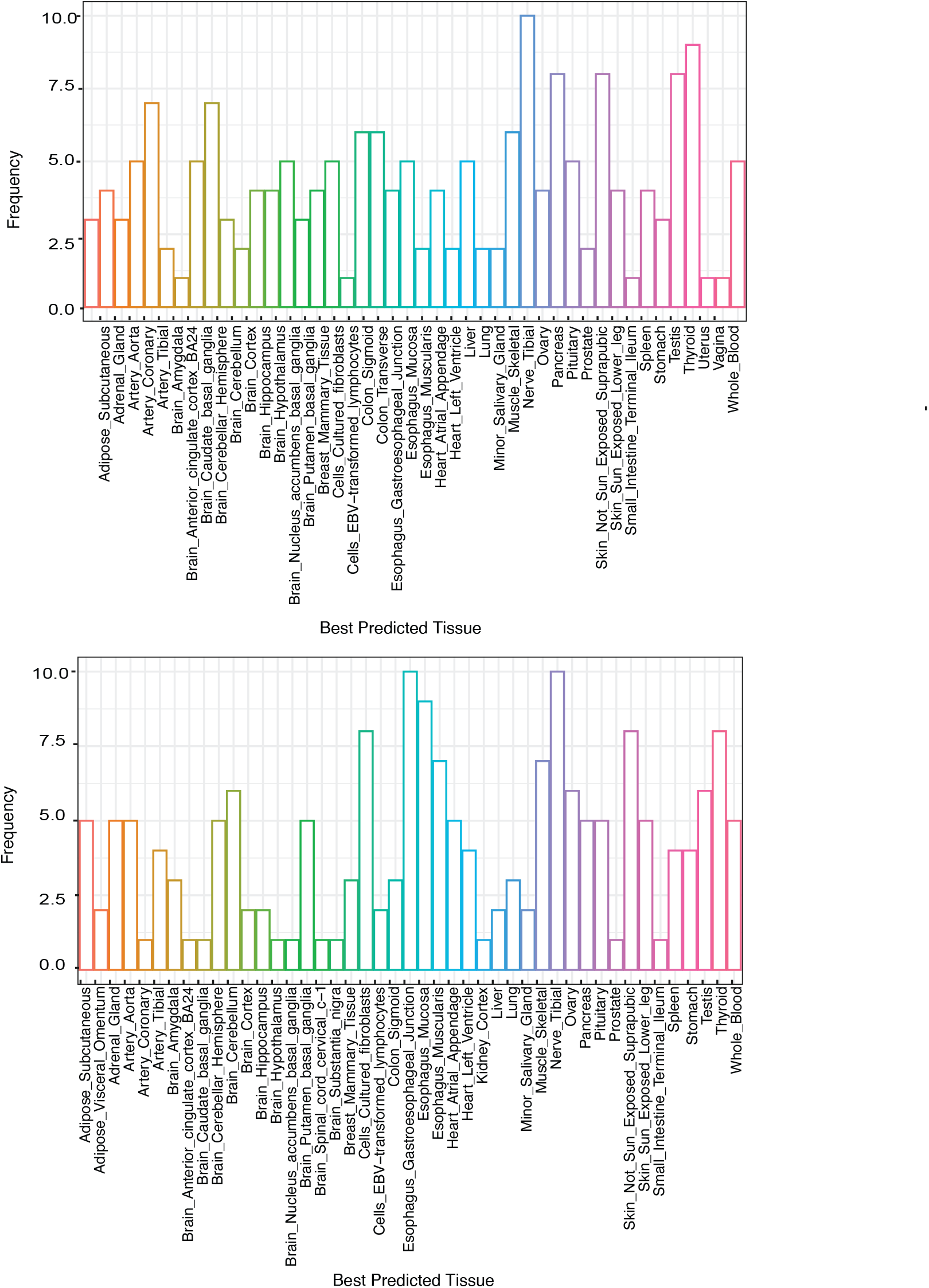
Best Predicted Tissue Distribution of Genes Associated between Depression and CAD. (a) Tissue distribution of Best Predicted Tissue for genes associated with both Depression and CAD using SMultiXcan Results of CAD (b) Tissue distribution of Best Predicted Tissue for genes associated with both Depression and CAD using SMultiXcan Results of Depression

